# Using fragmented data to characterize community healthcare utilization

**DOI:** 10.64898/2026.07.13.26357976

**Authors:** Taylor M. McCready, Lorna E. Thorpe, Brita Roy, Audrey Renson

## Abstract

Community-level estimates of healthcare utilization are essential for identifying inequities, allocating resources, and evaluating place-based interventions. However, in the United States, no single data source adequately captures healthcare utilization within geographically defined populations. Population-based surveys often lack sufficient geographic resolution, insurance claims represent only covered populations, and electronic health records are limited to care delivered within participating health systems. Increasingly, researchers combine these fragmented data sources, yet limited guidance exists for conducting valid population-based descriptive analyses using incomplete and overlapping data. We review the strengths and limitations of major data sources used to characterize community healthcare utilization and propose an approach for conducting population-based descriptive analyses using fragmented data. Rather than focusing on the limitations of individual data sources, our approach begins by explicitly defining the target population and the ideal observational study that would answer the research question. Available data sources are then conceptualized as incomplete or imperfect realizations of that ideal, providing a structured approach to (a) identifying sources of selection bias, missingness, and measurement error, (b) articulating required assumptions, and (c) selecting appropriate analytic strategies. We illustrate our approach using colorectal cancer screening utilization among adults residing in Brooklyn, New York during 2022. By shifting attention from individual data sources to the target community and the assumptions required for valid inference, this approach provides a practical approach for strengthening descriptive analyses of community healthcare utilization and informing place-based public health research, policy, and practice.

## INTRODUCTION

A common goal in public health is to characterize population-level patterns of healthcare utilization within geographically defined communities to target resources and address healthcare-related disparities. Community-level estimates of healthcare utilization help identify unmet healthcare needs, monitor inequities in access, allocate resources, and evaluate place-based interventions designed to improve population health. Policymakers and health systems increasingly seek to understand costs and outcomes of health encounters to optimize healthcare cost-effectiveness within a specific geographically-defined area, including assessing metrics that indicate missed opportunities for effective care, such as readmissions, non-emergent emergency department (ED) use,^1–9^ under-utilization of preventative services,^10^ and mismatches between healthcare needs and delivery.^11^ Given these questions are defined by where people live rather than where they receive care or who insures them, the target population often extends beyond the boundaries of any single healthcare system or insurance plan. Moreover, to further understand the promoters and barriers to care-seeking by underserved and minoritized populations, data on race/ethnicity, socioeconomic status, or other dimensions of social stratification are important ^12–14^ as a first step towards the identification of inequities and their associated upstream barriers and solutions.^15–17^

Obtaining representative estimates of healthcare utilization within communities remains challenging because no single data source adequately captures healthcare use across an entire population. Insurance claims and electronic health record (EHR) data are collected for administrative purposes, not research. As such, they represent only the subset of the geographic population that is covered by a given payer (claims) or captured within a given delivery system (EHR), rather than the full geographic population, and also suffer substantial measurement error and/or missing data. Conversely, public health surveys may be representative of certain populations but have limited geographic resolution and/or measurement of healthcare utilization. Increasingly, researchers seek to combine multiple data sources to overcome these limitations. However, combining fragmented data sources introduces additional challenges, including selection bias, inconsistent measurement across systems, incomplete follow-up, and uncertainty regarding the population represented by the combined data.

Limited guidance exists on how to thoughtfully conduct descriptive population-based analyses of healthcare utilization using multiple fragmented and overlapping disparate data sources.^18^ At the same time, the “data ecosystem” for utilization measurement has changed rapidly in recent years. During and after the COVID-19 era, health systems, public health agencies, and payers increasingly relied on near– real-time operational analytics, cross-institution data sharing, and centralized data platforms that can combine information from EHRs, laboratories, and claims. These developments have expanded what is feasible for measuring utilization within communities, but they also introduce new opportunities for selection bias, inconsistent capture across settings, and unclear denominators when data are assembled from multiple non-population-based sources.

While earlier reviews have been written on the use of healthcare utilization data,^19,20^ an update is warranted given rapid advancements in large health system data sources and emerging methodological guidance on ideal observational studies in recent years. The growth of EHRs and claims data, along with new centralized databases that join sources, offer new possibilities for comprehensive and updated analyses of healthcare utilization, along with additional challenges. ^18^ Existing reviews primarily describe the strengths and limitations of individual data sources but provide limited guidance for defining target populations, conceptualizing fragmented data relative to the research question, and selecting appropriate analytic strategies. This paper aims to fill this gap by providing a practical and targeted guide for researchers and practitioners engaged in descriptive population-based analyses of healthcare utilization, while considering possible limitations in available data sources. Our framework begins by defining the target population and the ideal observational study that would answer the research question, then conceptualizes available data sources as incomplete or imperfect realizations of that ideal. We illustrate the approach using a running example focused on colorectal cancer (CRC) screening utilization among adults aged 45–75 residing in Brooklyn, NY during calendar year 2022. The framework, however, is adaptable to other healthcare utilization questions that are either more or less broad in scope.

## A MOTIVATING EXAMPLE

To illustrate the type of questions of interest here, consider the following question: “*Among adults residing in Brooklyn, NY on January 1, 2022, what proportion were non-adherent to guideline-based CRC screening during calendar year 2022?”* Estimating community-level screening gaps in historically underserved neighborhoods can help identify unmet needs, target resources, and inform place-based interventions. This question is “population-based” in that the target population is defined by demographic and geographic factors rather than by membership in a particular health system or payer. This question is also descriptive in that it quantifies a utilization pattern and relevant covariates without attempting to estimate the effect of an intervention. While we focus on descriptive questions, the considerations discussed are also applicable to causal questions; however, causal analyses require additional considerations that are beyond the scope of this paper. We will return to our motivating question later when we discuss our proposed workflow.

### Data Sources for Characterizing Community Healthcare Utilization

Different data sources represent different segments of the target community and vary in how completely they capture healthcare utilization, leading to differences in the population represented and the assumptions required to make inferences about the broader community. We describe three types of data sources and their strengths and limitations for population-level health utilization studies: population-based surveys or disease registries, insurance claims, and EHRs.

### Population-Based Surveys and Disease Registries

Population-based surveys and disease registries have the advantage of being designed to represent defined populations rather than healthcare systems or insurance plans. For example, federally sponsored surveys such as the National Health Interview Survey (NHIS), Medical Expenditure Panel Survey (MEPS), and Behavioral Risk Factor Surveillance System (BRFSS), are designed to represent the US civilian noninstitutionalized population, and collect extensive data on demographics and health status.^21– 24^ However, surveys have many limitations. Geographic resolution is typically limited; NHIS enables national-level data estimates, while BRFSS allows for state and metro area inferences.^18,21,25,26^ Accessing finer geographic identifiers for some federal surveys (including NHIS) typically requires restricted-data approval and use of a Research Data Center environment, which can impose substantial time, cost, and output constraints. Even with approval, small sample sizes preclude estimation at fine geographic units, such as communities defined by zip code or census tract. Additionally, the main limitation of most surveys is that healthcare utilization data are self-reported. Accuracy of self-reported data varies greatly according to factors like recall period, visit type, and population characteristics,^27,28^ and agreement with administrative data has been observed to range between 30% and 99%.^27^ Despite these limitations, self-reported data can offer unique insights, particularly in capturing patient perspectives and experiences that are not always reflected in administrative records.

MEPS provides a key exception, as it links self-reported healthcare expenditure data to insurance claims.^23,29^ MEPS has many other strengths, including a longitudinal design; however, its limited sample size restricts analyses to the most common health conditions.^30^ In addition, MEPS public-use files are not designed for state or local estimation; the smallest geographic unit generally available is Census region, and smaller-area identifiers require restricted access (e.g., via the AHRQ Data Center/FSRDC process). Moreover, most public health surveys exclude or under-represent key groups (e.g., institutionalized, children, migrants).

Population-based disease registries such as the Surveillance, Epidemiology, and End Results (SEER) Program provide a census of disease cases within defined geographic populations (representing 48% of the U.S. population) and contain healthcare utilization data from administrative sources. ^31^ While SEER provides detailed geographic data down to the census-tract level, it includes limited demographic details at the individual level and focuses solely on disease-specific healthcare utilization.^24^

### Administrative Datasets

The main strengths of administrative datasets – including claims and EHR data – relative to surveys are that (i) utilization measures are more objective (though not without limitations), (ii) sample sizes are typically larger and can capture hard-to-reach subgroups, and (iii) geographic resolution is often finer. The major limitations are that administrative datasets typically comprise a highly-selected sample of the community, and suffer substantial, usually non-random, missing data.

#### Claims

Claims data (originating from healthcare providers’ invoices to insurers) track medical diagnoses, procedures, treatments, and prescriptions, along with costs and reimbursements.^32,33^ Because most utilization is billed, claims datasets capture nearly all utilization by members of the plans included. However, the representativeness of these datasets requires careful consideration and/or may be fundamentally uncertain. For example, commercial claims datasets represent individuals who purchased or have employer-provided insurance under a given plan or set of plans and are thus selected on many unobserved factors (e.g., occupation, income).

Public insurance claims can have clearer eligibility rules but still select community subgroups. Medicare covers most individuals over 65, those with disabilities, eligible children, and people with end stage renal disease, but Medicare claims datasets provided by CMS do not necessarily capture all claims for these individuals. In particular, there is a growing proportion (approximately 55% in 2026) of individuals who are also enrolled in a Medicare Part C Medicare Advantage (MA) plan offered by private insurers,^34^ and data on encounters/claims under MA may be incomplete or available through different access pathways than traditional fee-for-service Medicare claims. Thus, Medicare data can only be considered approximately representative of the Medicare-eligible population. However, an advantage is that it is rare for individuals to opt out of Medicare once enrolled, ensuring healthcare is tracked until the end of life.^34,35^

Medicaid claims represent individuals meeting income- and category-based eligibility criteria and therefore can be strongly selected with respect to socioeconomic status and other determinants of care-seeking. Medicaid datasets, which are available for some but not all states, include eligible low-income adults, children, pregnant women, elderly adults, and individuals with disabilities. ^36–38^ Geographic data for both Medicare and Medicaid are limited, generally allowing only national-level inferences. However, the Medicare Geographic Variation dataset allows for state and county-level analysis of health utilization, subject to privacy board approval.^39^

State all-payer claims databases (APCDs) can provide broader payer coverage within a community, but payer inclusion varies by state and may not fully capture public programs (including Medicaid) in all settings.^40^ Relatedly, many states maintain all-payer discharge data repositories, which contain summarized records for hospital admission and emergency department discharges, encompassing clinical details, diagnoses, performed procedures, duration of stay, and billing information. Additionally, a few states also track ambulatory surgery and outpatient services, providing a near-comprehensive view of utilization. However, discharge databases are limited to encounters occurring within reporting hospitals and typically do not capture longitudinal care across health systems or outpatient services unless those data are collected separately by the state.^41^ Zip code-level geographic markers are typically included, but demographic data are often limited to age and sex.^42^

Claims data present several methodological challenges, including measurement error and missing data, largely influenced by reimbursement-driven coding practices. Diagnoses and procedures are tracked via ICD and CPT codes, which are not intended for research, but are instead designed to facilitate the transfer of funds between payers and providers.^32^ Codes are sometimes selected for their higher reimbursement rates rather than for accurately representing the care reason.^32,43^ Additionally, there is variation in coding practices between settings (e.g., a family physician codes a diagnosis differently than a specialty physician)^44^ and over time, which can drive apparent trends in utilization. For example, apparent decreases in pneumonia hospitalization rates during the 2000s appear to be influenced by an increase in sepsis coding following a national sepsis awareness campaign.^45^ Additionally, changes to Medicare reimbursement rates can impact coding for Medicare and non-Medicare claims, as private insurance reimbursement rates often align with Medicare rates.^46^ Thus, to effectively work with claims data, researchers must consider policy or practice changes as well as shifts in public awareness that may impact coding patterns.

#### Electronic Health Records

Adoption of electronic health records has grown rapidly in recent years, enhancing the ability to aggregate documented healthcare utilization and outcomes; particularly for traditionally underserved populations, such as unhoused, uninsured, or migrant groups, and in geographically small areas.^47,48^ Though EHRs and claims both provide semi-objective measurement of utilization, EHRs often capture some utilization missed by claims; for instance, DeVoe et al. (2011) found that certain health screenings and tests were documented in EHRs but omitted in Medicaid claims (e.g., 7% of influenza vaccines).^49,50^ Similarly to claims, a major limitation of EHRs is that they only capture utilization within a single healthcare system or network of systems,^44^ and miss care received outside that network. Patient addresses are generally recorded and can be geocoded, but this requires restricted access, and not all systems maintain longitudinal records of historical patient addresses. Demographic data are typically available but may be limited due to high levels of missingness.^51^ For example, some systems (e.g., Epic) are increasingly capturing more detailed demographics such as education level and employment status, but there is a lack of unified standards across systems and over time,^52–54^ and some variables (e.g., race) may be missing or inaccurate.^55^

Multi-system EHR network datasets (e.g., TrinetX, Epic Cosmos) are increasingly available and have potential to reduce patient information fragmentation and enhance comprehensive health research. However, they come with challenges and have high potential for misuse, particularly with making broad inferences about patient care without consideration for the populations represented in these datasets. Some require patient consent to share data (e.g., health information exchanges), leading to highly selected samples.^56^ In addition, access may require health system participation in the network and can involve substantial contractual, governance, and per-user/per-project costs, which can limit feasibility and shape which communities and delivery systems are represented.

When multi-system EHR records are linked and harmonized at the individual level, variables collected in one system may partially “fill in” missingness in another—particularly for relatively stable attributes (e.g., race/ethnicity, language preference, education). However, linkage does not eliminate comparability concerns: data elements may be collected using different workflows (self-report vs staff-assigned), definitions, and value sets, and the same individual may have conflicting information across systems, creating uncertainty about the “true” value and requiring explicit reconciliation rules (e.g., source prioritization, most-recent value, or adjudication using self-report when available). Moreover, missingness and misclassification can remain systematically nonrandom if certain systems do not collect a field at all, collect it inconsistently over time, or do so with differential accuracy across patient groups. Finally, these networks may still incompletely capture utilization if major systems serving the community do not participate; patient matching errors and duplicate identities can introduce additional bias; and limited historical depth or delayed refresh cycles can constrain analyses of trends over time.

### REFINING THE SCOPE OF THE HEALTHCARE UTILIZATION ASSESSMENT

Analysis of healthcare utilization patterns in a defined population or geography begins by clearly specifying the scope of the question. Well-defined questions will state 1) the target population, 2) the time scale, 3) the time period, 4) the outcome, event or health state, and 5) the means of describing the outcome (e.g., prevalence). In specifying the question, one can begin by identifying the decisions it aims to inform; these components can then be defined to align with those decisions.

Time scale refers to how follow-up time is measured with reference to a particular time zero, which could be a person’s age, calendar time, or time since beginning treatment. Time period is the time lapse between time zero and the last outcome assessment. An ideal dataset typically contains data for every individual starting at time zero until the last observation (i.e., would have no late entry and no censoring).^57^ Depending on the outcome, complete ascertainment may also require information measured before time zero to establish eligibility or baseline outcome status. The outcome and summary measure should align with the public health or policy decision the analysis is intended to inform.

When tackling population-based descriptive questions about healthcare utilization with fragmented data, it can be helpful to imagine the ideal study that we would conduct to answer the question if resources and time were unlimited – i.e., fully representative, with perfect measurement of relevant variables at appropriate times, and no loss to follow-up. Although the ideal observational study is rarely feasible, describing the protocol of such a study can be a useful exercise because it requires sharpening aspects of the question which may otherwise remain obscure. Researchers can then identify how exactly the ideal study differs from the available data, delineate what assumptions need to be made to overcome these differences, plan analyses to approximate the answer from an ideal study under those assumptions, and transparently communicate remaining limitations. This approach builds on Jackson and colleagues’ “target study” framework, which specifies a hypothetical study protocol to guide the measurement of disparities using secondary data.^57^

We propose a simplified adaptation of this approach for descriptive analyses of healthcare utilization in which the goal is to estimate a utilization measure within a defined population rather than a disparity contrast between social groups. Accordingly, we specify the target population, eligibility criteria, time zero, follow-up, outcome assessment, and statistical analysis that would define the ideal observational study (Table 1). Components of the target study framework specific to disparity measurement, including enrollment groups and allowable covariates are not included.^57^

**Table 1.**
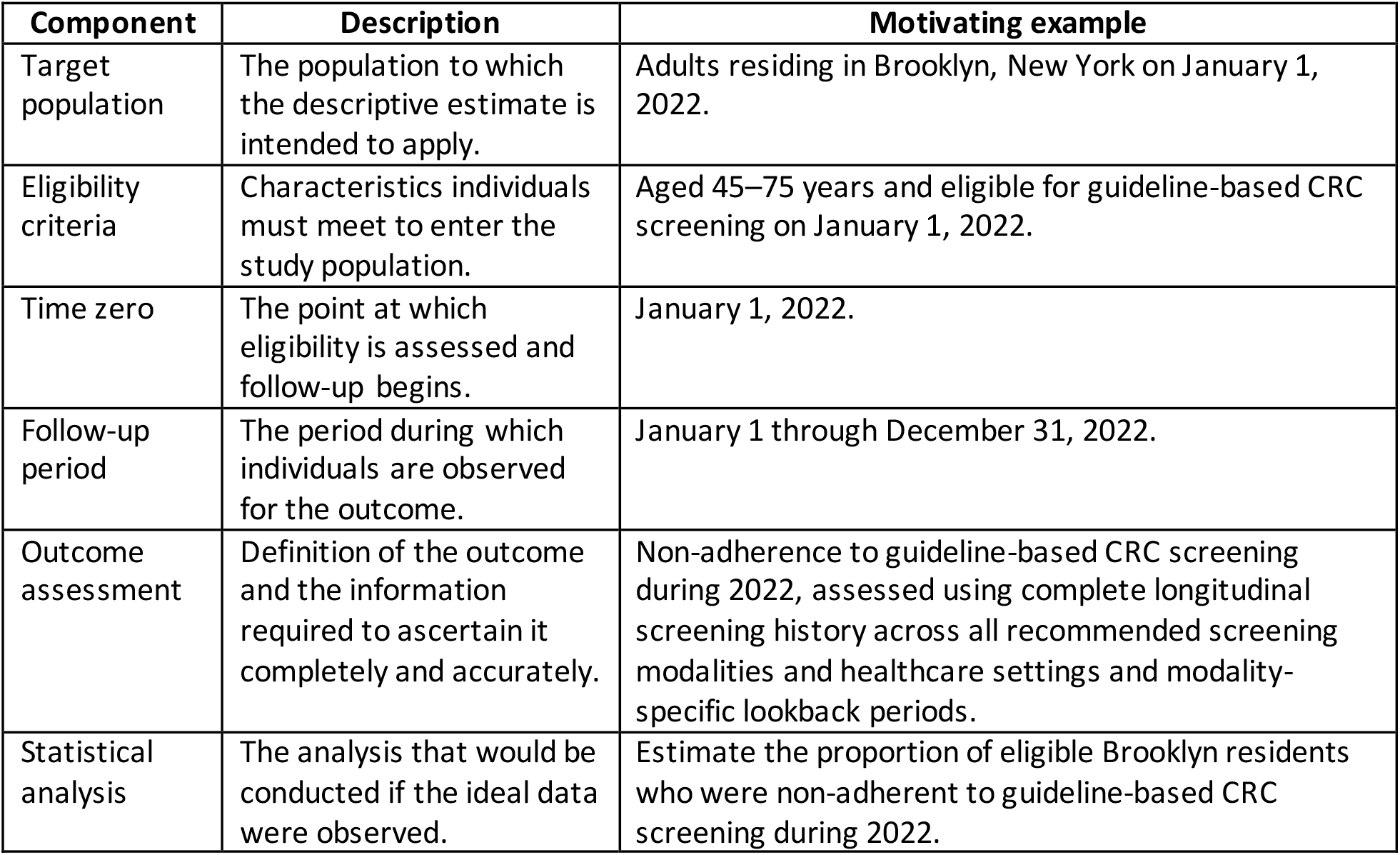
Specification of the ideal observational study for descriptive healthcare utilization analyses.

| **Component** | **Description** | **Motivating example** |
| --- | --- | --- |
| Target population | The population to which the descriptive estimate is intended to apply. | Adults residing in Brooklyn, New York on January 1, 2022. |
| Eligibility criteria | Characteristics individuals must meet to enter the study population. | Aged 45–75 years and eligible for guideline-based CRC screening on January 1, 2022. |
| Time zero | The point at which eligibility is assessed and follow-up begins. | January 1, 2022. |
| Follow-up period | The period during which individuals are observed for the outcome. | January 1 through December 31, 2022. |
| Outcome assessment | Definition of the outcome and the information required to ascertain it completely and accurately. | Non-adherence to guideline-based CRC screening during 2022, assessed using complete longitudinal screening history across all recommended screening modalities and healthcare settings and modality-specific lookback periods. |
| Statistical analysis | The analysis that would be conducted if the ideal data were observed. | Estimate the proportion of eligible Brooklyn residents who were non-adherent to guideline-based CRC screening during 2022. |

#### Emulating the ideal study

Once the ideal observational study has been defined, analysis proceeds by comparing available data with that ideal. This process involves three related steps. First, we identify how the observed data differ from the ideal study in terms of (i) the population or person-time represented and/or (ii) variables in terms of how and/or when they are measured. Second, we lay out what assumptions would be sufficient to equate a parameter estimated in the real dataset to the target summary parameter in the ideal study and decide whether these assumptions can be defended using substantive knowledge. Third, we conduct an analysis using those determined assumptions. In general, to address differences in population or person-time, we will typically need to assume that missingness is independent of the outcome given some set of covariates.^58^ Under such an assumption, we can use methods from the missing data and/or generalizability literature such as inverse probability of missingness weighting, multiple imputation, or doubly robust methods that combine the two. ^59^ To address measurement error, we typically need to assume that errors follow a known distribution conditional on observed data. Methods to correct for measurement error (e.g., multiple imputation) can then be employed.^59,60^

#### Example: Describing the ideal study

For our motivating example, the ideal observational study would include all adults aged 45–75 residing in Brooklyn on January 1, 2022 who were eligible for CRC screening based on recommendations of the US Preventive Services Task Force.^61^ Complete healthcare records would be available for every individual, allowing ascertainment of CRC screening history regardless of where care was received or the screening modality utilized (e.g., colonoscopy, sigmoidoscopy, or fecal immunochemical testing (FIT)). Given that recommended screening modalities have different screening intervals, accurately determining screening adherence requires longitudinal information preceding the study period.

In the ideal study, on the first of each month, we would record a variable for each individual that equals 1 if that individual was not up-to-date with screening as of that month (using the test-specific lookback window—e.g., up to 10 years for colonoscopy and shorter intervals for stool-based tests and endoscopic alternatives), and 0 otherwise. If an individual died or aged out of the relevant population as of that month, they would be no longer eligible to be non-adherent and receive a 0. At the end of one year, we would calculate a variable equal to 1 if each individual received a 1 for the non-adherence variable in any month, and 0 otherwise. Our relevant summary measure would be the mean of this variable – the proportion of individuals who were non-adherent during 2022, the denominator of which is the Brooklyn, NY population on January 1, 2022.

#### Emulating the ideal dataset

Either EHR or claims data likely represent the best hope of emulating the ideal dataset described above, since actual existing surveys lack small-area data granularity. One realistic scenario is that the research team might have access to a healthcare claims database that represents patients enrolled in managed care programs with value-based contracts in a large healthcare system serving the neighborhood of interest. But oftentimes other healthcare systems operate in that area as well. The analytic cohort would consist of eligible individuals represented in the claims database on January 1, 2022. Because recommended lookback periods differ across screening tests, the amount of historical information required to assess screening adherence varies across individuals.^62^ Some individuals may have incomplete historical information required to ascertain screening adherence at time zero because of prior changes in insurance coverage or enrollment in non-included plans. If an individual becomes uninsured or changes to a non-included plan during the study period, they are right-censored.

To emulate the ideal dataset, we must consider how and why our dataset differs, particularly regarding missing data. There are three sources of missing data relative to the ideal dataset: (i) claims data represent a non-random sample because some individuals may be insured by non-included plans, (ii) individuals may have incomplete historical information required to ascertain screening adherence at time zero, and (iii) individuals may have missing follow-up data (i.e., are right-censored). The latter two may occur because of changes in insurance or enrollment in non-included plans. In all three cases, individuals missing may differ systematically on variables that affect healthcare utilization, such as income, health-seeking behavior, age, sex, race/ethnicity, and immigration status. To emulate the ideal dataset, we need to account for such differences and fill in missing data using methods that require an assumption that CRC screening adherence is independent of missingness, conditional on measured covariates.

Of variables that might be required to meet that assumption, our claims dataset will likely only directly capture age, sex, and race/ethnicity along with payer type as a proxy for income. For health-seeking behavior, we could consider a proxy measure such as having a primary care visit in the last year. If we are willing to assume these are sufficient, analysis could then proceed by constructing inverse probability weights to standardize to the distribution of these variables as measured in the US decennial census and/or a large health survey such as the NYC Community Health Survey. For example, researchers could use post-stratification to align the age and sex distribution of the observed sample with that of the target population using population estimates from the American Community Survey.

We note that this approach contrasts with the common practice in claims data analyses of limiting the sample to those with continuous enrollment. Restricting to continuous enrollment has been observed to potentially introduce selection bias, regardless of the choice of enrollment period;^32,63,64^ thus, we do not advocate that approach here. Though our proposed approach can also incur biases if the assumptions are incorrect, defining a relevant time zero *a priori* and censoring individuals when they are lost to follow-up helps avoid selection bias by clearly defining the parameter of interest and the assumptions necessary to estimate it.

### Worked example

We illustrate this workflow using a multi-payer clinically integrated network (CIN) claims dataset. The target estimand is the proportion of adults aged 45–75 residing in Brooklyn (Kings County), NY on January 1, 2022 who were not up-to-date with guideline-based CRC screening at any point during calendar year 2022.

### Data structure and outcome definition

The CIN data contain repeated monthly measurements of a CRC screening care-gap indicator for attributed members. We defined the primary outcome as non-adherence during 2022, operationalized as an indicator equal to 1 if an individual was non-adherent with CRC screening in any month during 2022, and 0 otherwise.

### Early loss to follow-up as incomplete outcome ascertainment

Because individuals could become unattributed during 2022 (e.g., disenrollment from a participating payer), the outcome could not be completely ascertained for members who left the dataset before the end of 2022 and never had an observed open gap. We therefore treated early loss to follow-up as potentially informative right-censoring of the non-adherence outcome.

We summarized each individual’s monthly records and identified the last month observed in 2022. Individuals were classified as having complete outcome ascertainment (“fully observed”) if they either (1) were observed to be non-adherent during any month of 2022 (in which case the outcome was fully determined regardless of any subsequent unobserved months) or (2) remained attributed through the final measurement month of 2022 without being observed to be non-adherent. Individuals who were never observed to be non-adherent and became unattributed before the end of 2022 were classified as incompletely observed, because they could have become non-adherent after leaving the dataset. Incomplete ascertainment was thus one-sided: it could only obscure non-adherence, not adherence.

### Inverse probability-of-censoring weights

To adjust for differential loss to follow-up, we fit a logistic regression model for being fully observed as a function of baseline covariates available in the CIN data: age group (45–54, 55–64, 65–75 years), sex, preferred language group, payer type (commercial, Medicaid, Medicare), and census tract median household income quintile. Census tract income quintiles were defined using median household income estimates from the 2018–2022 American Community Survey (ACS) 5-year estimates for all census tracts in Brooklyn. We computed inverse probability-of-censoring weights (IPCW) as the inverse of each individual’s predicted probability of being fully observed. We examined the distribution of predicted probabilities of complete outcome ascertainment and the resulting IPCW to assess potential practical positivity violations and extreme weights.

### Post-stratification to the Brooklyn community age–sex distribution

To improve generalizability from the CIN-covered cohort to the Brooklyn community, we post-stratified the fully observed analytic sample to the joint age-by-sex distribution of Brooklyn residents aged 45–75 using person-level data from the 2018–2022 ACS 5-year Public Use Microdata Sample (PUMS). Single-year age was used to restrict the external target population to adults aged exactly 45–75 years. We constructed post-stratification weights for six age-by-sex strata defined by age group (45–54, 55–64, and 65–75 years) and sex. For each stratum, the post-stratification weight was calculated as the proportion of the PUMS target population in that stratum divided by the corresponding proportion of the fully observed CIN sample. We multiplied the post-stratification weights by the IPCW to obtain combined weights that addressed incomplete outcome ascertainment and differences in the age–sex distribution between the analytic sample and the Brooklyn population.

### Results of the worked example

The CIN-covered cohort included 16,428 adults aged 45–75. Among all 16,428 individuals, the observed proportion non-adherent at any point during 2022 was 41.1%. Of the 16,428 individuals, 1,322 (8.0%) left the dataset before the end of follow-up without a previously observed open care gap and were therefore classified as having incomplete outcome ascertainment. Among individuals with complete outcome ascertainment, applying IPCW yielded an estimated probability of non-adherence of 44.6%. After additionally applying age–sex post-stratification weights, the estimated probability of non-adherence at any point during 2022 was 45.7%.

### Assessing the plausibility of assumptions, interpretation, and limitations

The weighted estimate of CRC screening non-adherence should be interpreted as an estimate of the proportion of adults aged 45–75 residing in Brooklyn on January 1, 2022 who were non-adherent to guideline-based CRC screening during calendar year 2022 under the stated identifying assumptions. Rather than eliminating limitations inherent to fragmented healthcare data, the framework makes those limitations explicit by requiring researchers to define the target population, describe how available data differ from the ideal observational study, articulate the assumptions necessary to support the intended inference, and assess the plausibility of those assumptions.

In the worked example, we used a directed acyclic graph (DAG) to depict our assumptions about factors contributing to systematic differences between individuals represented in the CIN-covered cohort and the target population of Brooklyn residents (Figure 1). In the DAG shown in Figure 1, individual socioeconomic status is an unmeasured common cause of inclusion in the CIN-covered cohort and CRC screening non-adherence, with payer type and census tract socioeconomic characteristics serving as measured but imperfect proxies of individual socioeconomic status; payer type additionally determines attribution to the CIN directly. This structure highlights that weighting or standardization using measured characteristics may not fully account for selection into the available data if unmeasured socioeconomic differences remain associated with CRC screening. The DAG therefore makes explicit the assumptions underlying generalization to the target population and identifies potential sources of selection bias that should be considered when interpreting the standardized estimate and designing sensitivity analyses. We note that Figure 1 addresses selection into the CIN-covered cohort and generalization to the target population; assumptions regarding loss to follow-up within the cohort were discussed below. Finally, DAGs depict substantive assumptions rather than verify them; the validity of the analysis depends on whether the assumed structure adequately represents the underlying data-generating process.

**Figure 1.**
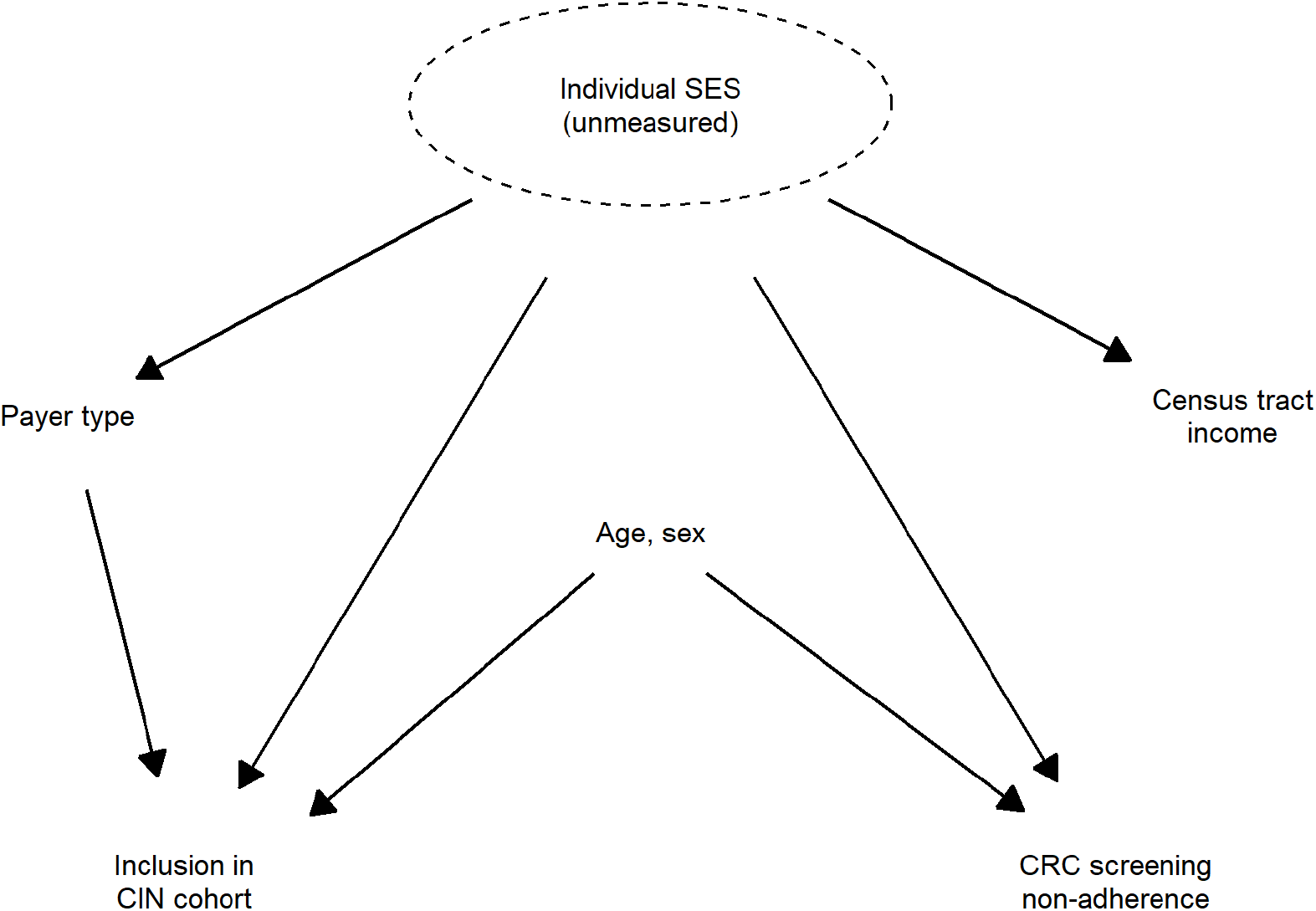
Directed acyclic graph depicts assumed sources of systematic difference between the CIN-covered cohort and the target population of adults aged 45–75 residing in Brooklyn, NY.

The IPCW analysis assumes that, among individuals never observed to be non-adherent, becoming unattributed before the end of 2022 was independent of subsequent non-adherence, conditional on the measured covariates in the censoring model. Equivalently, incompletely observed individuals are assumed to have the same conditional probability of non-adherence after loss to follow-up as comparable individuals who remained attributed and had not yet been observed non-adherent at the same point. This assumption may be violated because determinants of both continued observation and screening utilization, such as healthcare-seeking behavior, residential mobility, employment, insurance transitions, or use of healthcare systems outside the CIN, were incompletely measured or unavailable. Positivity is also required: within every covariate stratum of the censoring model represented in the CIN-covered cohort, individuals must have a nonzero probability of complete outcome ascertainment.

Predicted probabilities of complete outcome ascertainment were high and the resulting IPCW were stable, providing no evidence of severe practical positivity violations, although this does not establish the conditional exchangeability assumption.

Post-stratification additionally assumes that, within the age-by-sex strata used for standardization, the weighted CIN-covered population adequately represents the screening experience of the corresponding Brooklyn population. Standardization to the PUMS age–sex distribution addresses measured differences in these characteristics but cannot account for residual differences between the CIN-covered cohort and Brooklyn residents with respect to race and ethnicity, socioeconomic conditions beyond census tract income, insurance coverage, immigration-related factors, healthcare-seeking behavior, or other determinants of screening utilization. Post-stratification also requires that every age-by-sex stratum present in the Brooklyn target population be represented in the fully observed analytic sample; otherwise, the corresponding post-stratification weight would be undefined. All six strata were represented in the analytic sample. The 2018–2022 ACS 5-year PUMS also represents a pooled survey population rather than a census of Brooklyn residents on January 1, 2022 and should therefore be interpreted as an external standard for the age–sex distribution of the target community.

We examined whether the primary estimate was sensitive to two analytic decisions: the inclusion of census tract income quintile in the censoring model and the truncation of predicted probabilities used to construct the IPCW. Excluding census tract income quintile did not materially change the estimate, and alternative truncation specifications for the predicted probabilities used to construct the IPCW produced essentially identical estimates (45.7%). These analyses provide evidence that the primary estimate was not sensitive to these particular modeling decisions, but they do not assess sensitivity to unmeasured determinants of selection or incomplete outcome ascertainment. Quantitative bias analyses or other approaches that specify the magnitude of associations involving unmeasured factors could be used when sufficient substantive information is available to evaluate how departures from the identifying assumptions might affect the estimate.^65^ Although the motivating example uses claims data and CRC screening, the framework is not specific to either. The same principles can be applied to other healthcare utilization questions and to analyses based on electronic health records, surveys, disease registries, or linked data sources.

## Conclusion

Ultimately, the challenge is not that healthcare utilization data are fragmented, but that no available dataset perfectly represents the community we seek to understand. Beginning with the protocol of an ideal observational study provides a principled framework for clearly specifying the descriptive question. Ideally this process is done in collaboration with community stakeholders and/or other relevant decision-makers. Conceptualizing the available data as imperfect realizations of the ideal study, subject to missing data and/or measurement error, provides a principled approach to clarify the limitations of fragmented and imperfect healthcare data for describing communities. Assessing the plausibility of assumptions can also occur in a community engaged manner, for example through group model building. Together, these steps may help researchers generate more transparent, interpretable, and policy-relevant estimates of community healthcare utilization.

## Data Availability

All data produced in the present study are available upon reasonable request to the authors.

